# Regional Association of Disability and SARS-CoV-2 Infection in 369 Counties of the United States

**DOI:** 10.1101/2020.06.24.20139212

**Authors:** Oluwaseyi Olulana, Vida Abedi, Venkatesh Avula, Durgesh Chaudhary, Ayesha Khan, Shima Shahjouei, Jiang Li, Ramin Zand

## Abstract

**Background:** There have been outbreaks of SARS-CoV-2 in long term care facilities and recent reports of disproportionate death rates among the vulnerable population. The goal of this study was to better understand the impact of SARS-CoV-2 infection on the non-institutionalized disabled population in the United States using data from the most affected states as of April 9th, 2020.

**Methods:** This was an ecological study of county-level factors associated with the infection and mortality rate of SARS-CoV-2 in the non-institutionalized disabled population. We analyzed data from 369 counties from the most affected states (Michigan, New York, New Jersey, Pennsylvania, California, Louisiana, Massachusetts) in the United States using data available by April 9th, 2020. The variables include changes in mobility reported by Google, race/ethnicity, median income, education level, health insurance, and disability information from the United States Census Bureau. Bivariate regression analysis adjusted for state and median income was used to analyze the association between death rate and infection rate.

**Results:** The independent sample t-test of two groups (group 1: Death rate≥3.4% [median] and group 2: Death rate < 3.4%) indicates that counties with a higher total population, a lower percentage of Black males and females, higher median income, higher education, and lower percentage of disabled population have a lower rate (< 3.4%) of SARS-CoV-2 related mortality (all *p-*values<4.3E-02). The results of the bivariate regression when controlled for median income and state show counties with a higher White disabled population (est: 0.19, 95% CI: 0.01-0.37; *p-value*:3.7E-02), and higher population with independent living difficulty (est: 0.15, 95% CI: −0.01-0.30; *p-value*: 6.0E-02) have a higher rate of SARS-CoV-2 related mortality. Also, the regression analysis indicates that counties with higher White disabled population (est: - 0.22, 95% CI: −0.43-(-0.02); *p-*value: 3.3E-02), higher population with hearing disability (est: −0.26, 95% CI: - 0.42- (-0.11); *p*-value:1.2E-03), and higher population with disability in the 18-34 years age group (est: −0.25, 95% CI: −0.41-(-0.09); *p*-value:2.4E-03) show a lower rate of SARS-CoV-2 infection.

**Conclusion:** Our results indicate that while counties with a higher percentage of non-institutionalized disabled population, especially White disabled population, show a lower infection rate, they have a higher rate of SARS-CoV-2 related mortality.

## INTRODUCTION

The continuous spread of severe acute respiratory syndrome coronavirus 2 (SARS-CoV-2) has highlighted disparities in the infection and mortality rates of this disease.^1^ The outbreak in a nursing home in Kirkland, Washington^2^ was the first to report death due to SARS-CoV-2 in the United States (U.S). As of May 26, 2020, there are over 5.6 million confirmed cases worldwide, with over 1.6 million SARS-CoV-2 cases and 99,123 deaths in the U.S.^3^ SARS-CoV-2 initially attacks the respiratory system;^4^ with groups most susceptible to the infection and death being people aged 65 years and older, as well as people with certain chronic conditions and comorbidities. People living in nursing homes or long-term care facilities are more prone to the infection.^5,6^ Recently, states are releasing data on the number of deaths from long term care facilities^7^ with the most recent report stating that more than 10,000 patients and staff in long term facilities have died,^8^ accentuating the disparities in the outcomes of the disabled population in the U.S. The populations with higher disability are affected by the lower quality of care and increased comorbidities^9,10^ even before the evidence of disparities in the SARS-CoV-2 outcomes was reported. However, there have been few reports concerning the non-institutionalized disabled population and the risk of mortality and infection by SARS-CoV-2.^11^ To provide targeted preventive strategies, adequate resources for the disabled population and their care providers, an examination into the infection among the non-institutionalized disabled population is a necessary first step. The goal of this ecological study was to determine the association between county-level non-institutionalized disability rates, socioeconomic factors, and SARS-CoV-2 infection and death; we hypothesized that non-institutionalized disability rates in counties are associated with the rate of SARS-CoV-2 infection and death.

## METHODOLOGY

We conducted an ecological analysis of demographic, mobility, socioeconomic, and disability information, in the context of SARS-CoV-2 data in 369 counties in California, Michigan, New York, New Jersey, Louisiana, Pennsylvania, and Massachusetts with the initial highest SARS-CoV-2 infection and death rates as of April 9^th^, 2020.

### Data source

Data sources in this study include publicly available data from United States Census Bureau data estimated for 2018 for demographic data per county^12,13^ *USAFacts* for SARS-CoV-2 cases estimated for the year 2020,^14^ and mobility data provided by Google, as of April 5^th^, 2020.^15^

According to Social Security, individuals who cannot engage in substantial productive activity due to medically diagnosable physical or mental impairment which is expected to lead to death or last for over twelve months are legally defined as disabled.^16^ The non-institutionalized disabled population was defined as individuals not in a nursing home, prisons, or confined to any other facilities.

The data elements in this study included: 1) mobility data for each county, reported by Google on April 5^th^, 2020, that captured percent increase or decrease in mobility (grocery mobility, retail mobility, transit mobility, park mobility, work mobility, and resident mobility); 2) county-level data for total disabled population by race/ethnicity, age, gender, and type of disability along with data on total population, median income, education level, and access to health insurance, all data elements extracted from the United States Census Bureau. Table 1 summarizes a detailed description of the data elements used in this study. The outcome variables, infection and mortality rates were reported by USAFacts.^14^ County-level SARS-CoV-2 infection and mortality cases were extracted from the state’s health departments through data from hospitals, nursing homes, and other health organizations as reported on April 9^th^, 2020.

**Table 1.** Description of Data Elements.

| Data Elements | Description |
| --- | --- |
| Total population | Total population |
| SARS-CoV-2 infection rate | Rate of SARS-CoV-2 infection in one per million |
| SARS-CoV-2 infection death rate | Rate of SARS-CoV-2 infection related death (percent) |
| Retail mobility | Google retail recreation mobility |
| Grocery mobility | Google grocery pharmacy mobility |
| Park mobility | Google parks mobility |
| Transit mobility | Google transit mobility |
| Work mobility | Google workplace mobility |
| Resident mobility | Google residential mobility |
| Black male | Percent Black alone male |
| Black female | Percent Black alone female |
| Asian male | Percent Asian alone male |
| Asian female | Percent Asian alone female |
| White male | Percent Non-Hispanic White alone male |
| White female | Percent Non-Hispanic White alone female |
| Hispanic male | Percent Hispanic male |
| Hispanic female | Percent Hispanic female |
| Median income | Median family income; past 12 months 2018 |
| Poverty | Percent below poverty level population for whom poverty status is determined |
| High school | Percent below poverty level population for whom poverty status is determined RACE and HISPANIC OR LATINO ORIGIN White alone |
| Bachelors | Percent population 25 years and over bachelor's degree or higher |
| Insured male | Percent insured civilian noninstitutionalized population SEX male |
| Insured female | Percent insured civilian noninstitutionalized population SEX female |
| Insured Black | Percent insured civilian noninstitutionalized population RACE and HISPANIC OR LATINO ORIGIN Black or African American alone |
| Insured Asian | Percent insured civilian noninstitutionalized population RACE and HISPANIC OR LATINO ORIGIN ASIAN alone |
| Insured Hispanic | Percent insured civilian noninstitutionalized population RACE and HISPANIC OR LATINO ORIGIN Hispanic or Latino of any race |
| Insured White alone | Percent insured civilian noninstitutionalized population RACE and HISPANIC OR LATINO ORIGIN White alone not Hispanic or Latino |
| Medicaid | Percent public coverage Medicaid means tested public coverage alone or in combination |
| Disability | Percent with a disability total civilian non-institutionalized population |
| Disability male | Percent with a disability SEX Male |
| Disability female | Percent with a disability SEX Female |
| Disability Black | Percent with a disability RACE AND HISPANIC OR LATINO ORIGIN Black or African American |
| Disability Asian | Percent with a disability RACE AND HISPANIC OR LATINO ORIGIN Asian alone |
| Disability White | Percent with a disability RACE AND HISPANIC OR LATINO ORIGIN White alone, not Hispanic or Latino |
| Disability Hispanic | Percent with a disability RACE AND HISPANIC OR LATINO ORIGIN Hispanic or Latino (of any race) |
| Disability 5 to 17 years | Percent with a disability AGE 5 to 17 years |
| Disability 18 to 34 years | Percent with a disability AGE 18 to 34 years |
| Disability 35 to 64 years | Percent with a disability AGE 35 to 64 years |
| Disability 65 to 74 years | Percent with a disability AGE 65 to 74 years |
| Disability 75 years and over | Percent with a disability AGE 75 years |
| Disability hearing difficulty | Percent with a disability with a hearing difficulty |
| Disability vision difficulty | Percent with a disability with a vision difficulty |
| Disability cognitive difficulty | Percent with a disability with a cognitive difficulty |
| Disability ambulatory difficulty | Percent with disability with an ambulatory difficulty |
| Disability self-care difficulty | Percent with a disability with self-care difficulty |
| Disability independent living | Percent with a disability with independent living difficulty |

### Outcome Measures

The outcome measure is the county-level SARS-CoV-2 infection and mortality rates for the non-institutionalized disabled population. The infection and mortality rates were based on CDC guidelines^17^ (numerator: confirmed cases or death; denominator: total population per county). The mean of the infection rates or death rates were used for the comparative analysis as described below.

### Statistical Analysis

The data elements in this study were described using their mean ± standard deviation unless otherwise stated. An independent sample t-test was used to determine statistical significance in SARS-CoV-2 infection and death rates in comparison to the mobility, race/ethnicity, disability, median income, insured population, and total populations in the 369 counties. Bivariate regression adjusted for the state, and median income, ± total population was used to analyze the association between death rate or infection rate. Statistical analyses were performed using the IBM SPSS Statistics 26^18^ and R version 3.6.2.^19^

## RESULTS

### SARS-CoV-2 infection and mortality in the seven states during the first peak of infection in the U.S

The county-level data from 369 counties from Michigan, New York, New Jersey, Pennsylvania, Massachusetts, California, and Louisiana (mean population: 276,905.5±683,001.8) with the highest number of SARS-CoV-2 infection (as of April 9^th^, 2020) in the U.S. were included in this study.

The mean and 95% confidence interval (CI) for race/ethnicity, disability age group, and disability by gender are available in Table S1. The mean rate of disability in the U.S. population for the states used in the study was 15.0% ±3.7%; this rate for males was 13.1%±3.1% and for the female population was at 13.4%±2.8%. Disability in the race/ethnic groups ranged differently with the average of disability in the Black population (15.4% ±6.1%) as the highest; disability in the Asian population (7.9%±3.9%) was the lowest; disability rate in the white population was at 14.0%±2.9%. The age of the disabled population varied with the majority being 75 years or older (47.1%±6.0%) and lowest in disability age group 5 to 17 years at 6.3%±2.5%. Lastly, the disability types also varied within the population, ambulatory disability represents the highest type of disability at 7.0%±1.7% on average, while individuals with self-care difficulty were on average at 2.7%±0.7%. The hearing difficulty was at 3.7%±1.1%, while vision difficulty was at 2.2%±0.8%, cognitive difficulty at 5.4%±1.4%, and independent living disability was at 6.1%±1.4%. The mean infection rate was 1593.0±2768.9 per million with a median of 555.5, [IQR: 245.2-1486.6] while the mean mortality rate was 3.8%±2.2% and median of 3.4% [IQR: 2.2%-5.4%].

The independent sample t-test of the two groups (group 1: Death rate≥3.4% [median] and group 2: Death rate < 3.4%; Table 2) indicated that counties with a higher total population, reduction in retail mobility, a lower percentage of Black males and females, higher median income, higher education, lower percentage of disabled population, and a lower percentage of disabled in the age group of 35 to 64 years had a lower rate (< 3.4%) of SARS-CoV-2 related mortality (all *p-*values<4.3E-02). Additionally, a higher rate of SARS-CoV-2 infection was observed in counties with a higher population, a higher percentage of Black males and females, a higher median income, higher percentage of bachelor or graduate degree, a lower percentage of the disabled population, specifically a lower percentage of disabled individuals with hearing difficulty, and a lower percentage of disabled in the age group of 35 to 64 and 65 to 74 years (Table 3, all *p-*values<2.3E-02).

**Table 2.** Independent sample t-test comparing counties with a higher and lower median death rate.

| Data Elements | Total | | Group 1: SARS-CoV-2<br>death rate $\geq 3.4$ | | Group 2: SARS-CoV-2<br>death rate $< 3.4$ | | Significance |
| --- | --- | --- | --- | --- | --- | --- | --- |
|  | Mean<sup>+</sup> | Std.<br>Deviation | Mean<sup>+</sup> | Std.<br>Deviation | Mean<sup>+</sup> | Std.<br>Deviation |  |
| Total population | 276905.47 | 683001.80 | 295707.40 | 476086.20 | 605705.70 | 1113765.30 | 9.9E-03 |
| SARS-CoV-2 infection Rate* | 1593.00 | 2768.90 | 2483.40 | 3390.40 | 2306.30 | 3275.70 | 7.0E-01 |
| Retail mobility | -0.46 | 0.16 | -0.46 | 0.14 | -0.52 | 0.11 | 1.9E-03 |
| Grocery mobility | -0.16 | 0.15 | -0.16 | 0.13 | -0.20 | 0.14 | 3.3E-02 |
| Park mobility | 0.05 | 0.59 | 0.02 | 0.60 | 0.21 | 0.61 | 5.3E-02 |
| Transit mobility | -0.47 | 0.20 | -0.48 | 0.21 | -0.48 | 0.20 | 9.7E-01 |
| Work mobility | -0.37 | 0.10 | -0.38 | 0.09 | -0.40 | 0.07 | 7.5E-02 |
| Resident mobility | 0.14 | 0.06 | 0.15 | 0.06 | 0.14 | 0.04 | 8.1E-02 |
| Black male | 5.30 | 6.60 | 7.30 | 7.40 | 5.20 | 4.60 | 1.6E-02 |
| Black female | 4.90 | 6.90 | 7.30 | 8.00 | 5.40 | 5.30 | 4.3E-02 |
| Asian male | 1.50 | 2.50 | 1.65 | 2.35 | 2.80 | 3.50 | 6.2E-03 |
| Asian female | 1.60 | 2.70 | 1.80 | 2.50 | 3.10 | 3.80 | 5.8E-03 |
| White male | 37.10 | 9.90 | 34.00 | 10.70 | 33.30 | 9.50 | 6.3E-01 |
| White female | 37.60 | 10.00 | 34.70 | 10.80 | 34.70 | 10.80 | 9.4E-01 |
| Hispanic male | 5.20 | 6.70 | 6.00 | 8.10 | 7.20 | 6.90 | 2.3E-01 |
| Hispanic female | 4.80 | 6.60 | 5.80 | 8.10 | 7.00 | 6.80 | 2.4E-01 |
| Median income | 69910.10 | 19295.80 | 69778.60 | 19887.00 | 83430.40 | 20397.10 | 2.0E-06 |
| Poverty | 15.10 | 5.80 | 16.00 | 5.70 | 12.70 | 4.70 | 5.0E-06 |
| High school | 87.60 | 5.50 | 86.40 | 6.10 | 88.60 | 4.70 | 3.0E-03 |
| Bachelors | 24.70 | 10.80 | 24.50 | 10.80 | 31.80 | 10.90 | 2.0E-06 |
| Insured male | 91.50 | 3.40 | 91.10 | 3.20 | 92.50 | 3.00 | 1.5E-03 |
| Insured female | 93.60 | 3.00 | 93.20 | 3.10 | 94.40 | 2.50 | 2.8E-03 |
| Insured Black | 91.30 | 6.30 | 90.40 | 6.40 | 92.20 | 3.00 | 1.2E-02 |
| Insured Asian | 90.30 | 12.20 | 90.00 | 11.10 | 92.50 | 4.20 | 2.9E-02 |
| Insured Hispanic | 86.00 | 10.30 | 84.80 | 9.70 | 86.00 | 7.40 | 3.0E-01 |
| Insured White alone | 93.90 | 2.70 | 93.80 | 2.60 | 95.30 | 2.00 | 1.0E-05 |
| Medicaid | 23.20 | 6.60 | 24.30 | 7.30 | 20.60 | 6.40 | 1.7E-04 |
| Disability | 15.00 | 3.70 | 14.30 | 3.30 | 12.90 | 3.00 | 8.1E-04 |
| Disability male | 13.10 | 3.10 | 13.10 | 3.30 | 12.30 | 2.80 | 1.4E-01 |
| Disability female | 13.40 | 2.80 | 13.50 | 2.80 | 12.80 | 2.60 | 1.3E-01 |
| Disability Black | 15.40 | 6.10 | 15.50 | 4.70 | 14.80 | 6.40 | 4.9E-01 |
| Disability Asian | 7.90 | 3.90 | 7.30 | 3.00 | 7.80 | 4.10 | 5.2E-01 |
| Disability White | 14.00 | 2.85 | 14.20 | 3.10 | 13.30 | 2.50 | 5.0E-02 |
| Disability Hispanic | 11.40 | 4.90 | 11.70 | 6.10 | 11.10 | 4.20 | 5.2E-01 |
| Disability 5 to 17 years | 6.30 | 2.50 | 6.30 | 2.80 | 6.00 | 2.00 | 4.8E-01 |
| Disability 18 to 34 years | 7.10 | 2.30 | 7.20 | 2.30 | 6.70 | 2.20 | 1.4E-01 |
| Disability 35 to 64 years | 13.00 | 3.90 | 13.20 | 4.00 | 12.00 | 3.60 | 2.9E-02 |
| Disability 65 to 74 years | 23.30 | 5.20 | 23.60 | 5.50 | 22.60 | 5.00 | 2.4E-01 |
| Disability 75 years and over | 47.10 | 6.00 | 47.80 | 6.70 | 46.70 | 5.60 | 2.6E-01 |
| Disability hearing difficulty | 3.70 | 1.10 | 3.50 | 1.10 | 3.50 | 0.88 | 7.0E-01 |
| Disability vision difficulty | 2.20 | 0.77 | 2.30 | 0.89 | 2.10 | 0.67 | 1.5E-01 |
| Disability cognitive difficulty | 5.40 | 1.40 | 5.50 | 1.50 | 5.20 | 1.30 | 1.9E-01 |
| Disability ambulatory difficulty | 7.00 | 1.70 | 7.10 | 1.70 | 6.60 | 1.60 | 6.3E-02 |
| Disability self-care difficulty | 2.70 | 0.71 | 2.80 | 0.63 | 2.70 | 0.64 | 1.2E-01 |
| Disability independent living difficulty | 6.10 | 1.40 | 6.10 | 1.30 | 5.90 | 1.30 | 1.6E-01 |

+ All reported numbers are in percent except total population, SARS-CoV-2 infection rate, and median income.
\* Rate of SARS-CoV-2 infection in one per Million

**Table 3.** Independent sample t-test comparing counties with a higher and lower median infection rate.

| Data Elements | Total (N:352) | | Group 1: SARS-CoV-2 Infection Rate $\geq 555.6^*$ | | Group 2: SARS-CoV-2 Infection Rate $< 555.6^*$ | | Significance |
| --- | --- | --- | --- | --- | --- | --- | --- |
|  | Mean<sup>+</sup> | Std. Deviation | Mean<sup>+</sup> | Std. Deviation | Mean<sup>+</sup> | Std. Deviation |  |
| Total population | 276905.47 | 683001.80 | 382607.10 | 858300.10 | 196740.20 | 468817.40 | 1.2E-02 |
| SARS-CoV-2 death Rate | 3.83 | 2.20 | 3.50 | 2.00 | 4.60 | 2.60 | 5.0E-03 |
| Retail mobility | -0.46 | 0.16 | -0.49 | 0.15 | -0.44 | 0.15 | 3.0E-03 |
| Grocery mobility | -0.16 | 0.15 | -0.17 | 0.14 | -0.14 | 0.13 | 9.1E-02 |
| Park mobility | 0.05 | 0.59 | 0.24 | 0.52 | -0.20 | 0.57 | 6.4E-08 |
| Transit mobility | -0.47 | 0.20 | -0.47 | 0.21 | 0.47 | 0.17 | 8.6E-01 |
| Work mobility | -0.37 | 0.10 | -0.39 | 0.09 | -0.36 | 0.07 | 3.1E-02 |
| Resident mobility | 0.14 | 0.06 | 0.15 | 0.06 | 0.14 | 0.06 | 5.7E-01 |
| Black male | 5.30 | 6.60 | 8.20 | 7.30 | 2.60 | 4.30 | 1.8E-16 |
| Black female | 4.90 | 6.90 | 8.20 | 7.70 | 1.90 | 3.90 | 1.8E-19 |
| Asian male | 1.50 | 2.50 | 1.90 | 2.90 | 1.20 | 2.10 | 1.0E-02 |
| Asian female | 1.60 | 2.70 | 2.00 | 3.10 | 1.30 | 2.30 | 1.5E-02 |
| White male | 37.10 | 9.90 | 34.20 | 8.70 | 39.40 | 10.40 | 5.3E-07 |
| White female | 37.60 | 10.00 | 35.10 | 9.10 | 39.70 | 10.50 | 1.4E-05 |
| Hispanic male | 5.20 | 6.70 | 4.70 | 4.60 | 5.80 | 8.40 | 1.3E-01 |
| Hispanic female | 4.80 | 6.60 | 4.50 | 4.70 | 5.40 | 8.20 | 2.2E-01 |
| Median income | 69910.10 | 19295.80 | 76027.50 | 22996.80 | 65111.50 | 12876.50 | 8.9E-08 |
| Poverty | 15.10 | 5.80 | 14.90 | 6.30 | 15.30 | 5.10 | 5.1E-01 |
| High school | 87.60 | 5.50 | 87.40 | 5.40 | 87.60 | 5.60 | 7.9E-01 |
| Bachelors | 24.70 | 10.80 | 28.10 | 12.30 | 21.90 | 8.10 | 4.3E-08 |
| Insured male | 91.50 | 3.40 | 91.50 | 3.70 | 91.50 | 2.70 | 7.7E-01 |
| Insured female | 93.60 | 3.00 | 93.60 | 3.30 | 93.70 | 2.60 | 6.2E-01 |
| Insured Black | 91.30 | 6.30 | 90.70 | 5.50 | 91.80 | 6.90 | 9.7E-02 |
| Insured Asian | 90.30 | 12.20 | 90.00 | 13.10 | 90.10 | 11.50 | 9.5E-01 |
| Insured Hispanic | 86.00 | 10.30 | 84.20 | 10.60 | 87.50 | 9.10 | 2.7E-03 |
| Insured White alone | 93.90 | 2.70 | 94.20 | 2.90 | 93.70 | 2.30 | 1.0E-01 |
| Medicaid | 23.20 | 6.60 | 22.30 | 6.60 | 24.00 | 6.40 | 1.4E-02 |
| Disability | 15.00 | 3.70 | 14.10 | 3.70 | 15.60 | 3.40 | 2.3E-04 |
| Disability male | 13.10 | 3.10 | 12.60 | 3.10 | 13.80 | 3.00 | 6.2E-03 |
| Disability female | 13.40 | 2.80 | 13.10 | 2.80 | 13.80 | 2.80 | 8.6E-02 |
| Disability Black | 15.40 | 6.10 | 14.80 | 6.20 | 16.90 | 5.50 | 6.7E-02 |
| Disability Asian | 7.90 | 3.90 | 7.60 | 4.00 | 8.50 | 3.60 | 2.5E-01 |
| Disability White | 14.00 | 2.85 | 13.20 | 2.70 | 15.20 | 2.60 | 6.9E-07 |
| Disability Hispanic | 11.40 | 4.90 | 11.60 | 5.00 | 11.00 | 5.00 | 3.9E-01 |
| Disability 5 to 17 years | 6.30 | 2.50 | 6.40 | 2.40 | 6.20 | 2.70 | 6.7E-01 |
| Disability 18 to 34 years | 7.10 | 2.30 | 6.90 | 2.20 | 7.50 | 2.50 | 1.0E-01 |
| Disability 35 to 64 years | 13.00 | 3.90 | 12.50 | 4.10 | 13.70 | 3.50 | 2.3E-02 |
| Disability 65 to 74 years | 23.30 | 5.20 | 22.40 | 5.30 | 24.70 | 4.80 | 2.3E-03 |
| Disability 75 years and over | 47.10 | 6.00 | 46.60 | 6.20 | 47.70 | 5.60 | 1.8E-01 |
| Disability hearing difficulty | 3.70 | 1.10 | 3.40 | 0.96 | 4.10 | 1.10 | 9.0E-06 |
| Disability vision difficulty | 2.20 | 0.77 | 2.20 | 0.78 | 2.30 | 0.74 | 2.3E-01 |
| Disability cognitive difficulty | 5.40 | 1.40 | 5.30 | 1.50 | 5.50 | 1.30 | 3.0E-01 |
| Disability ambulatory difficulty | 7.00 | 1.70 | 6.80 | 1.70 | 7.20 | 1.60 | 1.3E-01 |
| Disability self-care difficulty | 2.70 | 0.71 | 2.80 | 0.70 | 6.00 | 1.40 | 8.3E-01 |
| Disability independent living difficulty | 6.10 | 1.40 | 6.00 | 1.40 | 6.20 | 1.30 | 4.2E-01 |

+ All reported numbers are in percent except total population, SARS-CoV-2 infection rate, and median income.
\* Rate of SARS-CoV-2 infection in one per Million

### Regression analysis showed a higher white disabled population, and independent living difficulty had a higher rate of SARS-CoV-2 related mortality

The results of the bivariate regression analysis (Figure 1, Table S2) estimate the effect size of disability sub-groups associated with SARS-CoV-2 related mortality when controlled for the median income and state. Counties with a higher percentage of White disabled population (est: 0.19, 95% CI: 0.01-0.37; *p-value*:3.7E-02), higher population with independent living difficulty (est: 0.15, 95% CI: −0.01-0.30; *p-value*: 6.0E-02), and higher disability in the age group 18-34 years (est:0.17, 95% CI: 0.02-0.31; *p*-value:2.4E-02) showed a higher rate of SARS-CoV-2 related mortality. The same trend is observed when controlling for the total population, median income and state in the bivariate regression model (Figure S1, Table S3).

**Figure 1.**
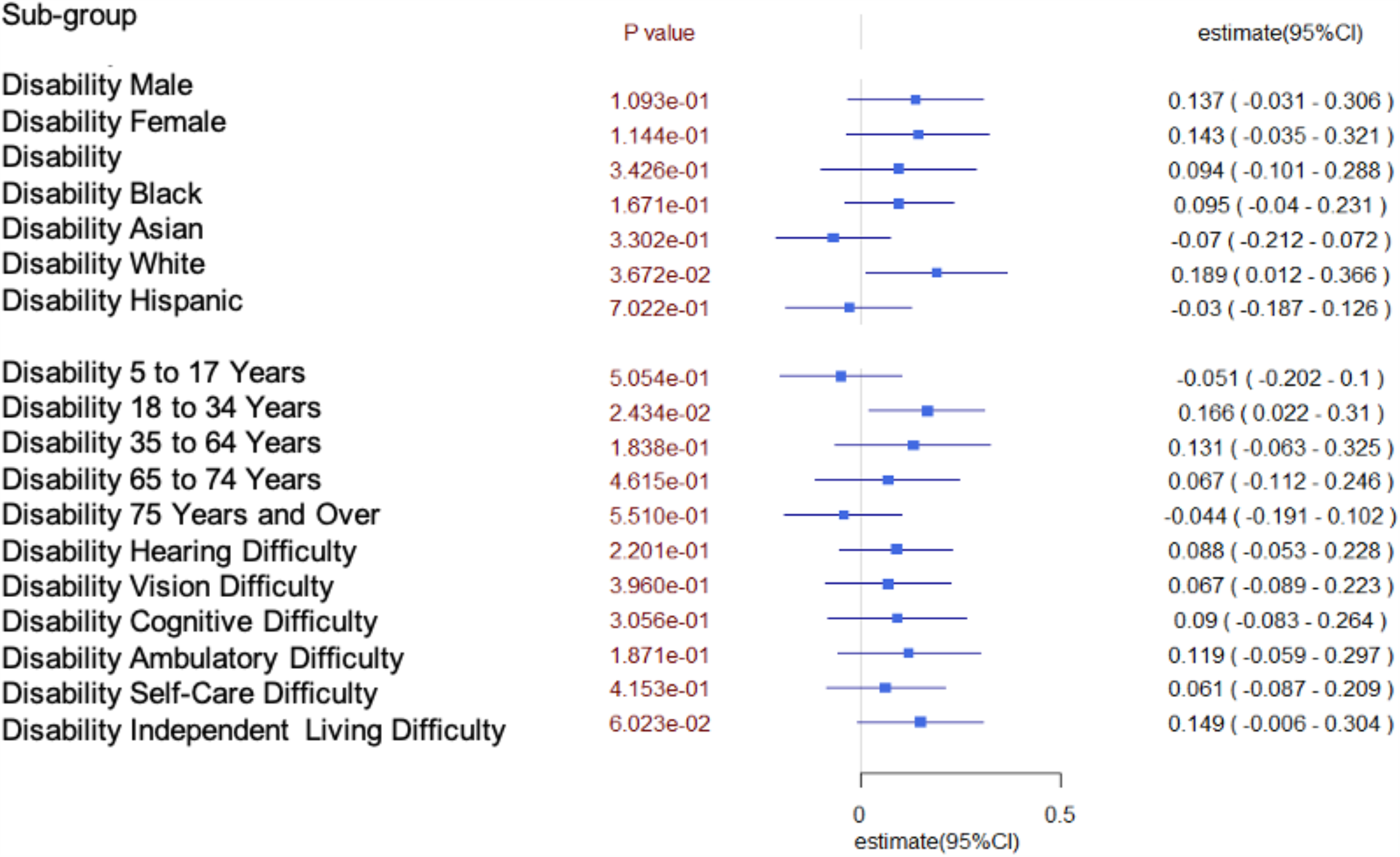
Death due to SARS-CoV-2 when controlled for state and median income.

### Regression analysis indicates no significant difference in SARS-CoV-2 related Infection rates among counties with different disability rates

The bivariate regression analysis was controlled for median income and state (Figure 2, Table S4), we observed that counties with higher percentage of White disabled population (est: −0.22, 95% CI: −0.43-(-0.02); *p-*value: 3.3E-02), higher percentage of people with hearing disability (est: −0.26, 95% CI: −0.42-(-0.11); *p*-value:1.2E-03), and higher percentage of people with disability in the 18-34 years age group (est: −0.25, 95% CI: −0.41-(-0.09); *p*-value:2.4E-03) showed a lower rate of SARS-CoV-2 infection. The bivariate regression analysis of the SARS-CoV-2 Infection rate when controlled for the total population, median income, and state showed no observed statistical significance in disability or any of the disability types (Figure S2, Table S5).

**Figure 2.**
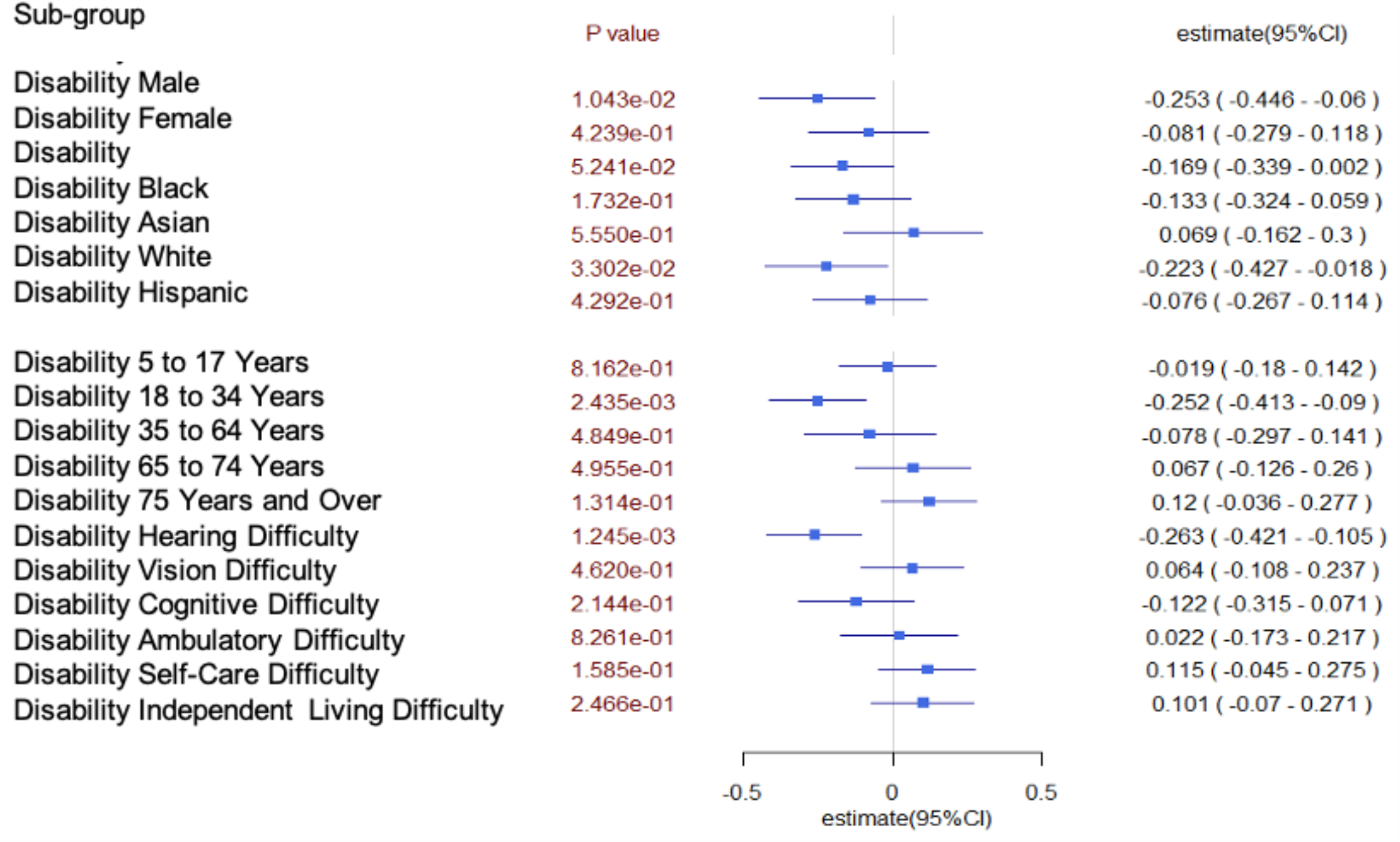
Infection due to SARS-CoV-2 when controlled for the state and median income.

## DISCUSSION

Our results indicate that counties with a higher percentage of non-institutionalized White disabled and a higher rate of independent living difficulty show a higher rate of SARS-CoV-2 related mortality. These results are plausible with the knowledge that the disabled population has a higher rate of obesity,^20–23^lack of physical activity,^20,22,23^ diabetes,^9,23,24^ higher rates of cardiovascular diseases^9,23,24^ that limits their body’s ability to fight the SARS-CoV-2 infection, which can ultimately lead to a poor outcome.^9,10^ Another reason for the high mortality rates could be due to the independent living difficulty that may have been compounded by the pandemic such as transportation limitations, grocery access, reduced opportunity for daily care, and essential services.^25^ Additionally, in anticipation of a supply crisis, many health departments around the U.S. considered options on how to triage patients who should be given the most aggressive life-saving therapies in the event of a supply shortage.^26^ In Washington state, a plan was put forward to withhold advanced care for patients with heart failure, chronic lung and liver disease among other organ dysfunctions; diseases that are common co-morbidities with the adult disabled population.^26–28^ Although this action was not officially taken in the U.S, there is a history of systematic discrimination in policies and treatment of disabled patients and lack of adequate preparedness for disabled patients during a pandemic.^23^

The center for disease control and prevention (CDC) cautioned that difficulty to understand information, limited mobility, and inability to communicate symptoms/illness may put the disabled patient population at risk.^29^ Alternatively, our results when adjusted for state and median income show counties with higher White disabled population and higher hearing disabled population have a lower rate of SARS-CoV-2 infection. There is a possibility that people with a hearing disability are overcompensated with visual information,^30^ which could potentially increase their attentiveness to the visual information concerning the spread of SARS-CoV-2 and their adherence to the enforced state-wide recommendations for the pandemic. Nonetheless, understanding how this sub-population is potentially better-protected needs further investigation.

Additionally, there was no significant difference in any of the disability groups when the analysis was controlled for population size, median income, and state. Although our bivariate regression analysis results on SARS-CoV-2 infection rate show there is no significance among states with different disability rates, this finding is not consistent with reports that the disabled population is at a higher risk of infection,^27,31^ we emphasize that the studied population was non-institutionalized disabled, so the trends may be different from studies that include both the institutionalized and non-institutionalized disabled population.^31^ The influenza pandemics showed that disabled patients are at an increased risk of infection, but also less likely to have access to vaccinations or be educated on their medical needs.^31^ The disabled population are at an increased risk of exposure, complication, and death due to their reliance on health services that may be on hold during pandemics, and if those services are available, they run the risk of exposure to the virus.^31^

This study provides some insight into how the counties with higher non-institutionalized disabled population may be affected by SARS-CoV-2 in the most affected states during the first wave of infection in the United States. SARS-CoV-2 is a call to pay more attention to this sub-population of non-institutionalized disabled civilians that are 12.7%^32^ of the total U.S population especially since the population is expected to see a 20% increase in adults over 65 years or older within the next decade,^33^ this age group is currently 40%^34^ of the disabled population. It is imperative that policies and programs are being put in place to avoid a public health crisis in the future for this group.^31^

Several limitations were observed in this study; the data used in this study was based on the county level. The data on the infection rate is likely an underestimate of the true cases, especially for the healthier population. In the future, we will continue to monitor and study the impact of the infection on this vulnerable population. In conclusion, our results indicate that while counties with a higher percentage of the non-institutionalized disabled population, especially White disabled population, show a lower infection rate, they have a higher rate of SARS-CoV-2 related mortality.

## Data Availability

The data that supports the findings of this school can be found on U.S Census, USAFacts and Google websites.

## Figure and Table Legends

**Table S1.** Independent sample t-test with comprehensive summary statistics comparing counties data element: mean, minimum, maximum, confidence interval lower and upper bound

| Data Elements | Mean<sup>+</sup> | Minimum | Maximum | 95% Confidence Interval Lower Bound | 95% Confidence Interval Upper Bound |
| --- | --- | --- | --- | --- | --- |
| Total population | 276905.5 | 1129.0 | 10039107.0 | 206987.7 | 346823.2 |
| SARS-CoV-2 infection rate* | 1593.0 | 15.4 | 20458.0 | 1302.8 | 1883.3 |
| SARS-CoV-2 related death rate | 3.8 | 0.3 | 10.1 | 3.5 | 4.1 |
| Retail mobility | -0.5 | -1.0 | 0.4 | -0.5 | -0.4 |
| Grocery mobility | -0.2 | -0.7 | 0.3 | -0.2 | -0.1 |
| Park mobility | 0.0 | -0.9 | 1.8 | 0.0 | 0.1 |
| Transit mobility | -0.5 | -0.9 | 0.7 | -0.5 | -0.4 |
| Work mobility | -0.4 | -0.7 | 0.5 | -0.4 | -0.4 |
| Resident mobility | 0.1 | 0.0 | 0.3 | 0.1 | 0.1 |
| Black male | 5.3 | 0.0 | 37.5 | 4.6 | 6.0 |
| Black female | 4.9 | 0.1 | 32.8 | 4.2 | 5.6 |
| Asian male | 1.5 | 0.0 | 19.0 | 1.2 | 1.7 |
| Asian female | 1.6 | 0.0 | 19.3 | 1.4 | 1.9 |
| White male | 37.1 | 4.5 | 50.3 | 36.1 | 38.1 |
| White female | 37.6 | 4.7 | 49.0 | 36.5 | 38.6 |
| Hispanic male | 5.2 | 0.4 | 42.3 | 72.6 | 76.7 |
| Hispanic female | 4.8 | 0.4 | 42.3 | 4.1 | 5.5 |
| Median income | 69910.1 | 30717.0 | 147878.0 | 67934.9 | 71885.4 |
| Poverty | 15.1 | 4.6 | 48.6 | 14.5 | 15.7 |
| High school | 87.6 | 68.5 | 96.1 | 87.0 | 88.1 |
| Bachelors | 24.6 | 7.4 | 60.8 | 23.5 | 25.7 |
| Insured male | 91.5 | 72.4 | 97.5 | 91.1 | 91.8 |
| Insured female | 93.6 | 80.1 | 98.4 | 93.3 | 93.9 |
| Insured Black | 91.4 | 46.7 | 100.0 | 90.7 | 92.0 |
| Insured Asian | 90.3 | 0.0 | 100.0 | 89.0 | 91.5 |
| Insured Hispanic | 85.9 | 30.3 | 100.0 | 84.9 | 87.0 |
| Insured White alone | 93.9 | 80.9 | 98.4 | 93.6 | 94.1 |
| Medicaid | 23.2 | 6.9 | 48.4 | 22.5 | 23.9 |
| Disability | 15.0 | 6.5 | 28.5 | 14.6 | 15.4 |
| Disability male | 13.1 | 7.1 | 22.3 | 12.6 | 13.5 |
| Disability female | 13.4 | 7.7 | 20.5 | 13.0 | 13.8 |
| Disability Black | 15.4 | 4.5 | 57.7 | 14.4 | 16.4 |
| Disability Asian | 7.9 | 2.3 | 33.1 | 7.1 | 8.6 |
| Disability White | 14.0 | 7.2 | 20.7 | 13.6 | 14.4 |
| Disability Hispanic | 11.4 | 3.6 | 33.0 | 10.6 | 12.2 |
| Disability 5 to 17 years | 6.3 | 1.9 | 13.9 | 6.0 | 6.7 |
| Disability 18 to 34 years | 7.1 | 2.3 | 16.4 | 6.8 | 7.4 |
| Disability 35 to 64 years | 13.0 | 4.9 | 25.6 | 12.5 | 13.6 |
| Disability 65 to 74 years | 23.3 | 10.0 | 36.1 | 22.6 | 24.1 |
| Disability 75 years and over | 47.1 | 32.3 | 67.7 | 46.2 | 47.9 |
| **Disability hearing difficulty** | 3.7 | 1.7 | 7.2 | 3.5 | 3.8 |
| **Disability vision difficulty** | 2.2 | 0.8 | 4.8 | 2.1 | 2.4 |
| **Disability cognitive difficulty** | 5.4 | 2.6 | 10.5 | 5.2 | 5.6 |
| **Disability ambulatory difficulty** | 7.0 | 3.4 | 11.2 | 6.7 | 7.2 |
| **Disability self-care Difficulty** | 2.7 | 1.1 | 6.1 | 2.6 | 2.8 |
| **Disability independent living difficulty** | 6.1 | 2.9 | 10.7 | 5.9 | 6.3 |

+ All reported numbers are in percent except total population, SARS-CoV-2 infection rate, and median income.
\* Rate of SARS-CoV-2 infection in one per Million

**Table S2.** Detailed results of the bivariate analysis of risk of death by SARS-CoV-2 when controlled for state, and median income

| Risk Factors | Estimate | Standard Error | Statistic | p-value | Conf.low | Conf.high | Estimate_95CI |
| --- | --- | --- | --- | --- | --- | --- | --- |
| Disability | 9.40E-02 | 9.87E-02 | 9.51E-01 | 3.43E-01 | -1.01E-01 | 2.88E-01 | 0.09 (-0.10 - 0.29) |
| Disability male | 1.37E-01 | 8.52E-02 | 1.61E+00 | 1.09E-01 | -3.11E-02 | 3.06E-01 | 0.14 (-0.03 - 0.31) |
| Disability female | 1.43E-01 | 9.00E-02 | 1.59E+00 | 1.14E-01 | -3.49E-02 | 3.21E-01 | 0.14 ( -0.04 - 0.32) |
| Disability Black | 9.50E-02 | 6.85E-02 | 1.39E+00 | 1.67E-01 | -4.04E-02 | 2.31E-01 | 0.10 ( -0.04 - 0.23) |
| Disability Asian | -7.00E-02 | 7.13E-02 | -9.79E-01 | 3.30E-01 | -2.12E-01 | 7.19E-02 | -0.07 ( -0.21 - 0.07) |
| Disability White alone | 1.89E-01 | 8.98E-02 | 2.11E+00 | 3.67E-02 | 1.18E-02 | 3.66E-01 | 0.19 (0.012 - 0.37) |
| Disability Hispanic | -3.00E-02 | 7.92E-02 | -3.83E-01 | 7.02E-01 | -1.87E-01 | 1.26E-01 | -0.03 (-0.19 - 0.13) |
| Disability 5 to 17 Years | -5.10E-02 | 7.63E-02 | -6.67E-01 | 5.05E-01 | -2.02E-01 | 9.99E-02 | -0.05(-0.20 - 0.1) |
| Disability 18 to 34 Years | 1.66E-01 | 7.30E-02 | 2.27E+00 | 2.43E-02 | 2.18E-02 | 3.10E-01 | 0.17 (0.02 - 0.31) |
| Disability 35 to 64 Years | 1.31E-01 | 9.81E-02 | 1.33E+00 | 1.84E-01 | -6.28E-02 | 3.25E-01 | 0.13 (-0.06 - 0.33) |
| Disability 65 to 74 Years | 6.70E-02 | 9.06E-02 | 7.38E-01 | 4.62E-01 | -1.12E-01 | 2.46E-01 | 0.07 ( -0.11 - 0.25) |
| Disability 75 Years and Over | -4.40E-02 | 7.41E-02 | -5.98E-01 | 5.51E-01 | -1.91E-01 | 1.02E-01 | -0.04 (-0.191 - 0.10) |
| Disability hearing difficulty | 8.80E-02 | 7.12E-02 | 1.23E+00 | 2.20E-01 | -5.30E-02 | 2.28E-01 | 0.09 (-0.05 - 0.23) |
| Disability vision difficulty | 6.70E-02 | 7.88E-02 | 8.51E-01 | 3.96E-01 | -8.86E-02 | 2.23E-01 | 0.07 (-0.09 - 0.22) |
| Disability cognitive difficulty | 9.00E-02 | 8.79E-02 | 1.03E+00 | 3.06E-01 | -8.33E-02 | 2.64E-01 | 0.09 (-0.08 - 0.26) |
| Disability ambulatory difficulty | 1.19E-01 | 9.00E-02 | 1.32E+00 | 1.87E-01 | -5.85E-02 | 2.97E-01 | 0.12 (-0.06 - 0.30) |
| Disability self-care difficulty | 6.10E-02 | 7.48E-02 | 8.17E-01 | 4.15E-01 | -8.67E-02 | 2.09E-01 | 0.06 (-0.09 - 0.21) |
| Disability independent living difficulty | 1.49E-01 | 7.85E-02 | 1.89E+00 | 6.02E-02 | -6.47E-03 | 3.04E-01 | 0.15 ( -0.01 - 0.30) |

**Table S3.** Detailed results of the bivariate analysis of the risk of death by SARS-CoV-2 when controlled for the state, median income, and total population

| Risk Factors | Estimate | Standard Error | Statistic | p-value | Conf.low | Conf.high | Estimate_95CI |
| --- | --- | --- | --- | --- | --- | --- | --- |
| **Disability** | 5.30E-02 | 1.05E-01 | 5.03E-01 | 6.16E-01 | -1.54E-01 | 2.59E-01 | 0.05 ( -0.15 - 0.26) |
| **Disability male** | 1.59E-01 | 9.97E-02 | 1.60E+00 | 1.12E-01 | -3.75E-02 | 3.56E-01 | 0.16 ( -0.04 - 0.36) |
| **Disability female** | 1.48E-01 | 9.73E-02 | 1.52E+00 | 1.31E-01 | -4.46E-02 | 3.40E-01 | 0.15 ( -0.05 - 0.34) |
| **Disability Black** | 1.11E-01 | 7.28E-02 | 1.53E+00 | 1.30E-01 | -3.31E-02 | 2.55E-01 | 0.11 ( -0.03 - 0.26) |
| **Disability Asian** | -6.00E-02 | 7.56E-02 | -7.90E-01 | 4.32E-01 | -2.10E-01 | 9.07E-02 | -0.06 ( -0.21 - 0.09) |
| **Disability White alone** | 2.08E-01 | 9.93E-02 | 2.10E+00 | 3.77E-02 | 1.20E-02 | 4.05E-01 | 0.21 (0.012 - 0.41) |
| **Disability Hispanic** | -3.30E-02 | 8.39E-02 | -3.91E-01 | 6.96E-01 | -1.99E-01 | 1.33E-01 | -0.03 ( -0.20 - 0.13) |
| **Disability 5 to 17 Years** | -5.70E-02 | 8.11E-02 | -7.03E-01 | 4.83E-01 | -2.17E-01 | 1.03E-01 | -0.06 ( -0.22 - 0.10) |
| **Disability 18 to 34 Years** | 1.95E-01 | 8.19E-02 | 2.38E+00 | 1.89E-02 | 3.27E-02 | 3.57E-01 | 0.20 (0.03 - 0.36) |
| **Disability 35 to 64 Years** | 1.46E-01 | 1.06E-01 | 1.37E+00 | 1.72E-01 | -6.43E-02 | 3.56E-01 | 0.15 ( -0.06 - 0.36) |
| **Disability 65 to 74 Years** | 7.50E-02 | 9.46E-02 | 7.91E-01 | 4.30E-01 | -1.12E-01 | 2.62E-01 | 0.08 ( -0.11 - 0.26) |
| **Disability 75 Years and Over** | -4.50E-02 | 7.65E-02 | -5.88E-01 | 5.57E-01 | -1.96E-01 | 1.06E-01 | -0.05 ( -0.20 - 0.11) |
| **Disability hearing difficulty** | 9.00E-02 | 8.58E-02 | 1.05E+00 | 2.97E-01 | -7.99E-02 | 2.59E-01 | 0.09 ( -0.08 - 0.26) |
| **Disability vision difficulty** | 7.40E-02 | 8.21E-02 | 8.99E-01 | 3.70E-01 | -8.84E-02 | 2.36E-01 | 0.07 ( -0.09 - 0.24) |
| **Disability cognitive difficulty** | 1.04E-01 | 9.48E-02 | 1.10E+00 | 2.73E-01 | -8.30E-02 | 2.92E-01 | 0.10 ( -0.08 - 0.30) |
| **Disability ambulatory difficulty** | 1.17E-01 | 9.51E-02 | 1.23E+00 | 2.21E-01 | -7.12E-02 | 3.05E-01 | 0.12 ( -0.07 - 0.31) |
| **Disability self-care difficulty** | 7.70E-02 | 7.82E-02 | 9.84E-01 | 3.27E-01 | -7.76E-02 | 2.32E-01 | 0.08 ( -0.08 - 0.23) |
| **Disability independent living difficulty** | 1.62E-01 | 8.21E-02 | 1.97E+00 | 5.04E-02 | -2.82E-04 | 3.24E-01 | 0.16 (0 - 0.32) |

**Table S4.** Detailed results of the bivariate analysis of the risk of infection by SARS-CoV-2 when controlled for state, and median income

| Risk Factors | Estimate | Standard error | Statistic | p-value | Conf.low | Conf.high | Estimate_95CI |
| --- | --- | --- | --- | --- | --- | --- | --- |
| Disability | -1.69E-01 | 8.64E-02 | -1.95E+00 | 5.24E-02 | -3.39E-01 | 1.78E-03 | -0.17 (-0.34 - 0.002) |
| Disability male | -2.53E-01 | 9.75E-02 | -2.60E+00 | 1.04E-02 | -4.46E-01 | -6.03E-02 | -0.25 (-0.45 - -0.06) |
| Disability female | -8.10E-02 | 1.00E-01 | -8.02E-01 | 4.24E-01 | -2.79E-01 | 1.18E-01 | -0.08 (-0.28 - 0.12) |
| Disability Black | -1.33E-01 | 9.67E-02 | -1.37E+00 | 1.73E-01 | -3.24E-01 | 5.91E-02 | -0.13 ( -0.32 - 0.06) |
| Disability Asian | 6.90E-02 | 1.16E-01 | 5.93E-01 | 5.55E-01 | -1.62E-01 | 3.00E-01 | 0.07 (-0.16 - 0.3) |
| Disability White alone | -2.23E-01 | 1.03E-01 | -2.15E+00 | 3.30E-02 | -4.27E-01 | -1.82E-02 | -0.22 ( -0.43 - -0.02) |
| Disability Hispanic | -7.60E-02 | 9.63E-02 | -7.93E-01 | 4.29E-01 | -2.67E-01 | 1.14E-01 | -0.08 ( -0.27 - 0.11) |
| Disability 5 to 17 Years | -1.90E-02 | 8.14E-02 | -2.33E-01 | 8.16E-01 | -1.80E-01 | 1.42E-01 | -0.02 (-0.18 - 0.14) |
| Disability 18 to 34 Years | -2.52E-01 | 8.16E-02 | -3.09E+00 | 2.44E-03 | -4.13E-01 | -9.05E-02 | -0.25( -0.41 - -0.09) |
| Disability 35 to 64 Years | -7.80E-02 | 1.11E-01 | -7.00E-01 | 4.85E-01 | -2.97E-01 | 1.41E-01 | -0.08 ( -0.30 - 0.14) |
| Disability 65 to 74 Years | 6.70E-02 | 9.76E-02 | 6.83E-01 | 4.96E-01 | -1.26E-01 | 2.60E-01 | 0.07 ( -0.13 - 0.26) |
| Disability 75 Years and Over | 1.20E-01 | 7.94E-02 | 1.52E+00 | 1.31E-01 | -3.65E-02 | 2.77E-01 | 0.12 ( -0.04 - 0.28) |
| Disability hearing difficulty | -2.63E-01 | 7.99E-02 | -3.29E+00 | 1.25E-03 | -4.21E-01 | -1.05E-01 | -0.26 ( -0.42 - -0.11) |
| Disability vision difficulty | 6.40E-02 | 8.72E-02 | 7.38E-01 | 4.62E-01 | -1.08E-01 | 2.37E-01 | 0.06 (-0.108 - 0.24) |
| Disability cognitive difficulty | -1.22E-01 | 9.78E-02 | -1.25E+00 | 2.14E-01 | -3.15E-01 | 7.13E-02 | -0.12 ( -0.32 - 0.07) |
| Disability ambulatory difficulty | 2.20E-02 | 9.86E-02 | 2.20E-01 | 8.26E-01 | -1.73E-01 | 2.17E-01 | 0.02 ( -0.17 - 0.22) |
| Disability self-care difficulty | 1.15E-01 | 8.10E-02 | 1.42E+00 | 1.59E-01 | -4.53E-02 | 2.75E-01 | 0.12 ( -0.05 - 0.28) |
| Disability independent living difficulty | 1.01E-01 | 8.64E-02 | 1.16E+00 | 2.47E-01 | -7.03E-02 | 2.71E-01 | 0.10 ( -0.07 - 0.27) |

**Table S5.** Detailed results of the bivariate analysis of the rate of infection by SARS-CoV-2 when controlled for the state, median income, and total population

| Risk Factors | Estimate | Standard Error | Statistic | p-value | Conf.low | Conf.high | Estimate_95CI |
| --- | --- | --- | --- | --- | --- | --- | --- |
| Disability | -8.40E-02 | 8.27E-02 | -1.02E+00 | 3.11E-01 | -2.47E-01 | 7.91E-02 | -0.08 (-0.25 - 0.08) |
| Disability male | -8.90E-02 | 9.70E-02 | -9.21E-01 | 3.59E-01 | -2.81E-01 | 1.02E-01 | -0.09 (-0.28 - 0.10) |
| Disability female | -1.60E-02 | 9.34E-02 | -1.72E-01 | 8.64E-01 | -2.01E-01 | 1.69E-01 | -0.02 (-0.20 - 0.17) |
| Disability Black | -1.14E-01 | 8.94E-02 | -1.28E+00 | 2.04E-01 | -2.92E-01 | 6.32E-02 | -0.11 (-0.29 - 0.06) |
| Disability Asian | 1.40E-01 | 1.09E-01 | 1.28E+00 | 2.05E-01 | -7.81E-02 | 3.58E-01 | 0.14 (-0.08 - 0.36) |
| Disability White alone | -5.80E-02 | 1.01E-01 | -5.70E-01 | 5.69E-01 | -2.58E-01 | 1.43E-01 | -0.06 (-0.26 - 0.14) |
| Disability Hispanic | -7.00E-03 | 8.94E-02 | -8.28E-02 | 9.34E-01 | -1.85E-01 | 1.70E-01 | -0.01 (-0.19 - 0.17) |
| Disability 5 to 17 Years | 7.80E-02 | 7.46E-02 | 1.04E+00 | 2.99E-01 | -6.98E-02 | 2.25E-01 | 0.08 (-0.07 - 0.23) |
| Disability 18 to 34 Years | -1.40E-01 | 7.84E-02 | -1.78E+00 | 7.71E-02 | -2.95E-01 | 1.54E-02 | -0.14 (-0.30 - 0.02) |
| Disability 35 to 64 Years | -6.30E-02 | 1.02E-01 | -6.13E-01 | 5.41E-01 | -2.65E-01 | 1.40E-01 | -0.06 (-0.265 - 0.14) |
| Disability 65 to 74 Years | -1.10E-02 | 8.94E-02 | -1.18E-01 | 9.06E-01 | -1.87E-01 | 1.66E-01 | -0.01 (-0.19 - 0.17) |
| Disability 75 Years and Over | 7.10E-02 | 7.25E-02 | 9.82E-01 | 3.28E-01 | -7.22E-02 | 2.15E-01 | 0.07 (-0.07 - 0.22) |
| Disability hearing difficulty | -8.80E-02 | 8.40E-02 | -1.05E+00 | 2.97E-01 | -2.54E-01 | 7.82E-02 | -0.09 (-0.25 - 0.08) |
| Disability vision difficulty | 1.20E-02 | 7.96E-02 | 1.46E-01 | 8.84E-01 | -1.46E-01 | 1.69E-01 | 0.01 (-0.15 - 0.17) |
| Disability cognitive difficulty | -7.00E-02 | 8.96E-02 | -7.83E-01 | 4.35E-01 | -2.47E-01 | 1.07E-01 | -0.07 (-0.25 - 0.11) |
| Disability ambulatory difficulty | 3.50E-02 | 9.06E-02 | 3.87E-01 | 7.00E-01 | -1.44E-01 | 2.14E-01 | 0.04 ( -0.14 - 0.21) |
| Disability self-care difficulty | 4.90E-02 | 7.47E-02 | 6.52E-01 | 5.15E-01 | -9.91E-02 | 1.97E-01 | 0.05 (-0.10 - 0.20) |
| Disability independent living difficulty | 6.50E-02 | 7.89E-02 | 8.30E-01 | 4.08E-01 | -9.05E-02 | 2.21E-01 | 0.07 (-0.091 - 0.22) |

**Figure S1.**
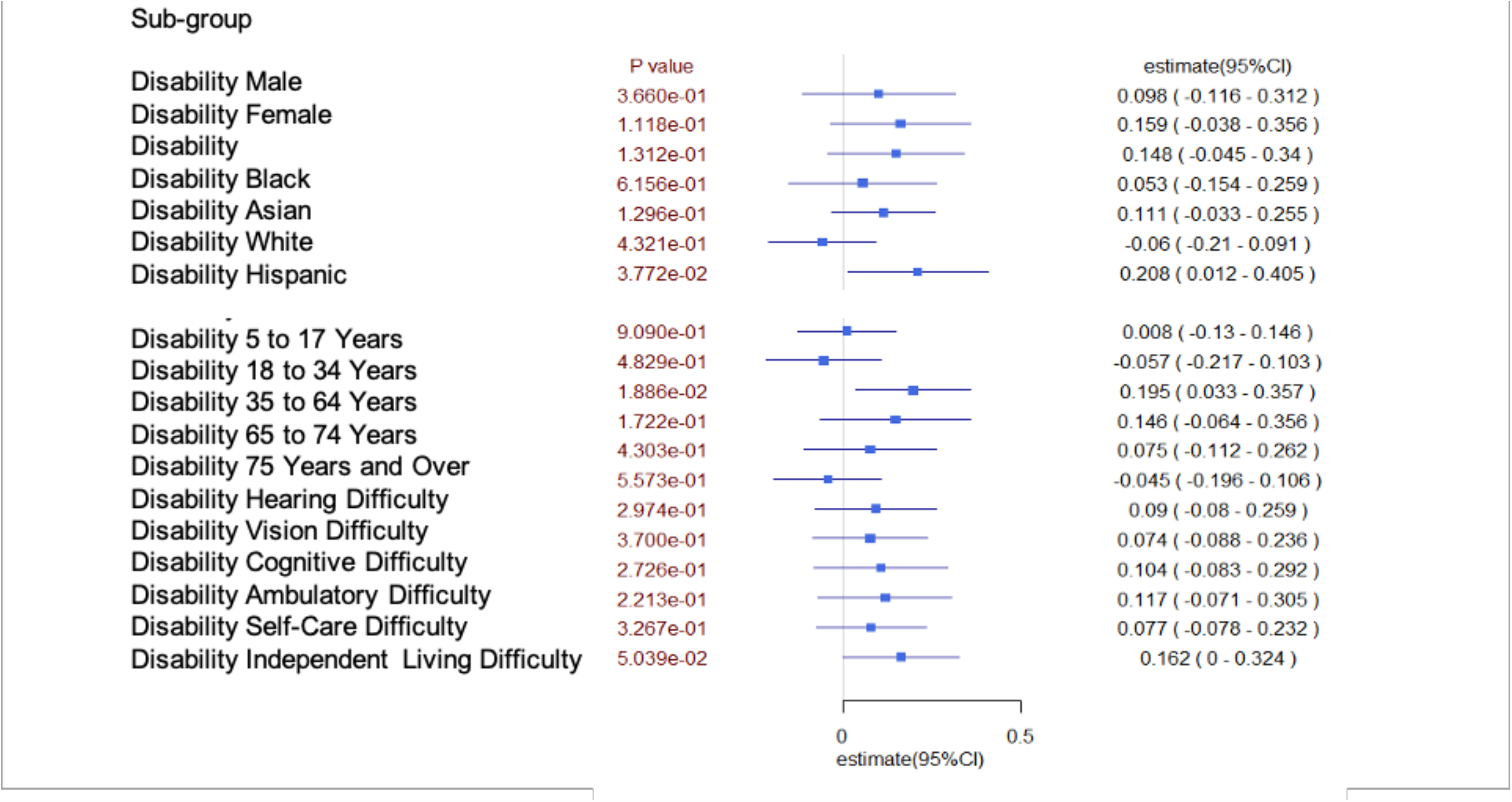
Death due to SARS-CoV-2 controlled for state, median income and total population.

**Figure S2.**
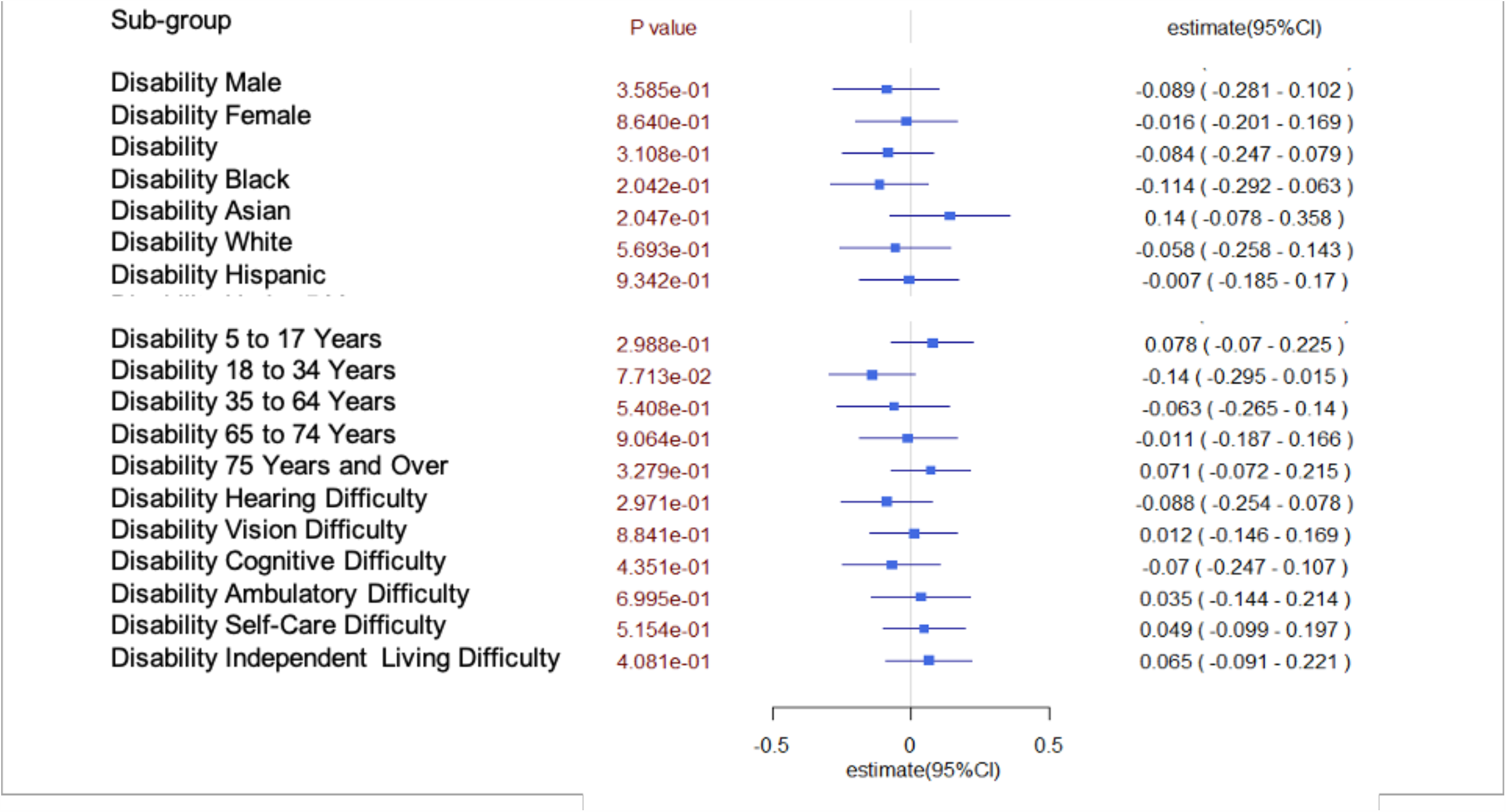
Infection due to SARS-CoV-2 controlled for the state, median income and total population.

